# The NYCKidSeq randomized controlled trial: Impact of GUÍA digitally enhanced genetic counseling in racially and ethnically diverse families

**DOI:** 10.1101/2023.07.05.23292193

**Authors:** Sabrina A. Suckiel, Nicole R. Kelly, Jacqueline A. Odgis, Katie M. Gallagher, Monisha Sebastin, Katherine E. Bonini, Priya N. Marathe, Kaitlyn Brown, Miranda Di Biase, Michelle A. Ramos, Jessica E. Rodriguez, Laura Scarimbolo, Beverly J. Insel, Kathleen D.M. Ferar, Randi E. Zinberg, George A. Diaz, John M. Greally, Noura S. Abul-Husn, Laurie J. Bauman, Bruce D. Gelb, Carol R. Horowitz, Melissa P. Wasserstein, Eimear E. Kenny

**Affiliations:** Institute for Genomic Health, Icahn School of Medicine at Mount Sinai, New York, NY; Department of Medicine, Icahn School of Medicine at Mount Sinai, New York, NY; Department of Pediatrics, Division of Pediatric Genetic Medicine, Children’s Hospital at Montefiore/Montefiore Medical Center/Albert Einstein College of Medicine, Bronx, NY; Department of Population Health Science and Policy, Icahn School of Medicine at Mount Sinai, New York, NY; Institute for Health Equity Research, Icahn School of Medicine at Mount Sinai, New York, NY; Department of Genetics and Genomic Sciences, Icahn School of Medicine at Mount Sinai, New York, NY; Department of Obstetrics, Gynecology and Reproductive Science, Icahn School of Medicine at Mount Sinai, New York, NY; Department of Pediatrics, Icahn School of Medicine at Mount Sinai, New York, New York; Department of Psychiatry and Behavioral Sciences, Albert Einstein College of Medicine, Bronx, NY; Department of Pediatrics, Albert Einstein College of Medicine, Bronx, NY; Mindich Child Health and Development Institute, Icahn School of Medicine at Mount Sinai, New York, NY

**Keywords:** digital tools, digital solutions, genetic testing, diverse populations, genetic counseling, genomic medicine

## Abstract

**Background:** Digital solutions are needed to support rapid increases in the application of genetic and genomic tests (GT) in diverse clinical settings and patient populations. We developed GUÍA, a bi-lingual web-based platform that facilitates disclosure of GT results. The NYCKidSeq randomized controlled trial evaluated GUÍA’s impact on understanding of GT results.

**Methods:** NYCKidSeq enrolled diverse children with neurologic, cardiac, and immunologic conditions who underwent GT. Families were randomized to genetic counseling with GUÍA (intervention) or standard of care (SOC) genetic counseling for results disclosure. Parents/legal guardians (participants) completed surveys at baseline, post-results disclosure, and 6-months later. Survey measures assessed the primary study outcomes of perceived understanding of and confidence in explaining their child’s GT results and the secondary outcome of objective understanding. We used regression models to evaluate the association between the intervention and the study outcomes.

**Results:** The analysis included 551 participants, 270 in the GUÍA arm and 281 in SOC. Participants’ mean age was 41.1 years and 88.6% were mothers. Most participants were Hispanic/Latino(a) (46.3%), White/European American (24.5%), or Black/African American (15.8%). Participants in the GUÍA arm had significantly higher perceived understanding post-results (OR=2.8, CI[1.004,7.617], *P*=0.049) and maintained higher objective understanding over time (OR=1.1, CI[1.004, 1.127], *P*=0.038) compared to those in the SOC arm. There was no impact on perceived confidence. Hispanic/Latino(a) individuals in the GUÍA arm maintained higher perceived understanding (OR=3.9, CI[1.6, 9.3], *P*=0.003), confidence (OR=2.7, CI[1.021, 7.277], *P*=0.046), and objective understanding (OR=1.1, CI[1.009, 1.212], *P*=0.032) compared to SOC.

**Conclusions:** This trial demonstrates that GUÍA positively impacts understanding of GT results in diverse parents of children with suspected genetic conditions. These findings build a case for utilizing GUÍA to deliver complex and often ambiguous genetic results. Continued development and evaluation of digital applications in diverse populations are critical for equitably scaling GT offerings in specialty clinics.

## Introduction

Rapid advancements in genomic sequencing over the last two decades have resulted in an increasing number of patients receiving results from genetic and genomic tests (GT) in various clinical settings. This growing availability of GT, coupled with technological innovation in digital technology and Artificial Intelligence (AI), has resulted in an increasing convergence between healthcare and technology toward enhancing and scaling genomic medicine. A vital component of genomic medicine is the communication of GT results to patients and their families, which is usually carried out by genetic counselors via in-person or telehealth counseling. This type of effective, patient-centered communication is essential for patients to understand their results, and across medical research, studies have found that understanding test results can positively impact adherence to medical follow-up, overall satisfaction, and psychological adaptation(1,2). While the positive impact of genetic counseling on patients’ understanding of their GT results is well established(3), relying solely on current practices as genome medicine becomes more prevalent in health systems is likely impractical given an insufficient genetics workforce and rapidly expanding patient populations receiving GT results(4).

Digital solutions are emerging as valuable tools for expanding access and supporting the GT process(5,6). For example, decision support tools use web- or app-based interfaces to help patients and families decide whether or not to undergo GT(7,8), make decisions about medical management related to their genomic risk(9), and choose whether to receive secondary findings from genomic sequencing(10). Patients can access self-guided educational material and modules(11–13), which help to streamline pre-test counseling. Chatbots are also helpful in facilitating the genetic counseling and testing process by providing an AI-driven mechanism for collecting family history, delivering education, performing risk assessments, and helping patients share results with family members(14–17). Typically, digital genetic counseling tools address the more routine aspects of the pre-test counseling workflow, such as family history collection and education, since these are often simpler to standardize. Far fewer applications are focused on communicating GT results (post-test counseling)(18). Communicating GT results to patients can be challenging due to the complexity of the results, the degree of technical language used in clinical GT reports, the potential for misinterpretation, and the range of result types from benign to uncertain to pathogenic(19,20). Digital tools could potentially improve the communication of complex genetic results with features such as digital post-test educational modules, personalization, and visualization of results and medical recommendations. Additionally, digital tools could be designed to support better patients with low health literacy or limited English proficiency to understand and potentially act on highly technical GT information.

To investigate whether digital tools designed for results disclosure genetic counseling could improve the understanding of GT results, we created a web-based application called GUÍA (Genomic Understanding, Information, and Awareness application), which has been previously described(21). GUÍA, which means ‘guide’ in Spanish, is designed to facilitate the communication of GT results to patients and families. The application encompasses relevant aspects of a results disclosure genetic counseling session, including genetic education, primary (related to indication for testing) and secondary (unrelated to the indication for testing) results disclosure, clinical implications, inheritance and family implications, medical recommendations, and patient resources. Providers can personalize their patient’s application by inputting the genetic results and other relevant information into the software. GUÍA uses text, images, and a modular design to help convey information comprehensively. It also enables patients to actively engage with their results and control the speed and depth of information delivery. Importantly, GUÍA was developed with input from community stakeholders committed to ensuring that the application was suitable for diverse populations, is written at a ninth-grade reading level, and is currently available in English and Spanish.

Here, we report the findings from NYCKidSeq, a randomized controlled trial (RCT) jointly-funded by the National Human Genome Research Institute and the National Institute on Minority Health and Health Disparities, and one of seven national clinical sites that are part of the Clinical Sequencing Evidence-Generating Research (CSER) consortium(22). The NYCKidSeq RCT was designed to compare digitally enhanced (using GUÍA) genetic counseling to traditional genetic counseling in results disclosure sessions of diverse pediatric patients and their families recruited from two large health systems in New York City(23). The objectives of the study were to assess the impact of GUÍA on parents’ perceived understanding of and confidence in explaining their child’s GT results (primary outcomes) and on objective understanding of the GT results (secondary outcome). Evaluating the effectiveness of GUÍA in NYCKidSeq sheds light on the acceptability and utility of using this digital application to support the return of GT results to families of pediatric patients.

## Methods

### Study Design

The NYCKidSeq study design, described previously in Odgis *et al*.(23), is an RCT (Clinicaltrials.gov: NCT03738098) in which pediatric children received GT and were randomized to one of two study arms to receive either digitally enhanced (using GUÍA) genetic counseling or traditional genetic counseling (standard of care) (see **Figure 1**). The Institutional Review Boards at the Icahn School of Medicine at Mount Sinai (IRB# 17-00780) and the Albert Einstein College of Medicine (IRB# 2016-6883) approved the study.

**Figure 1.**
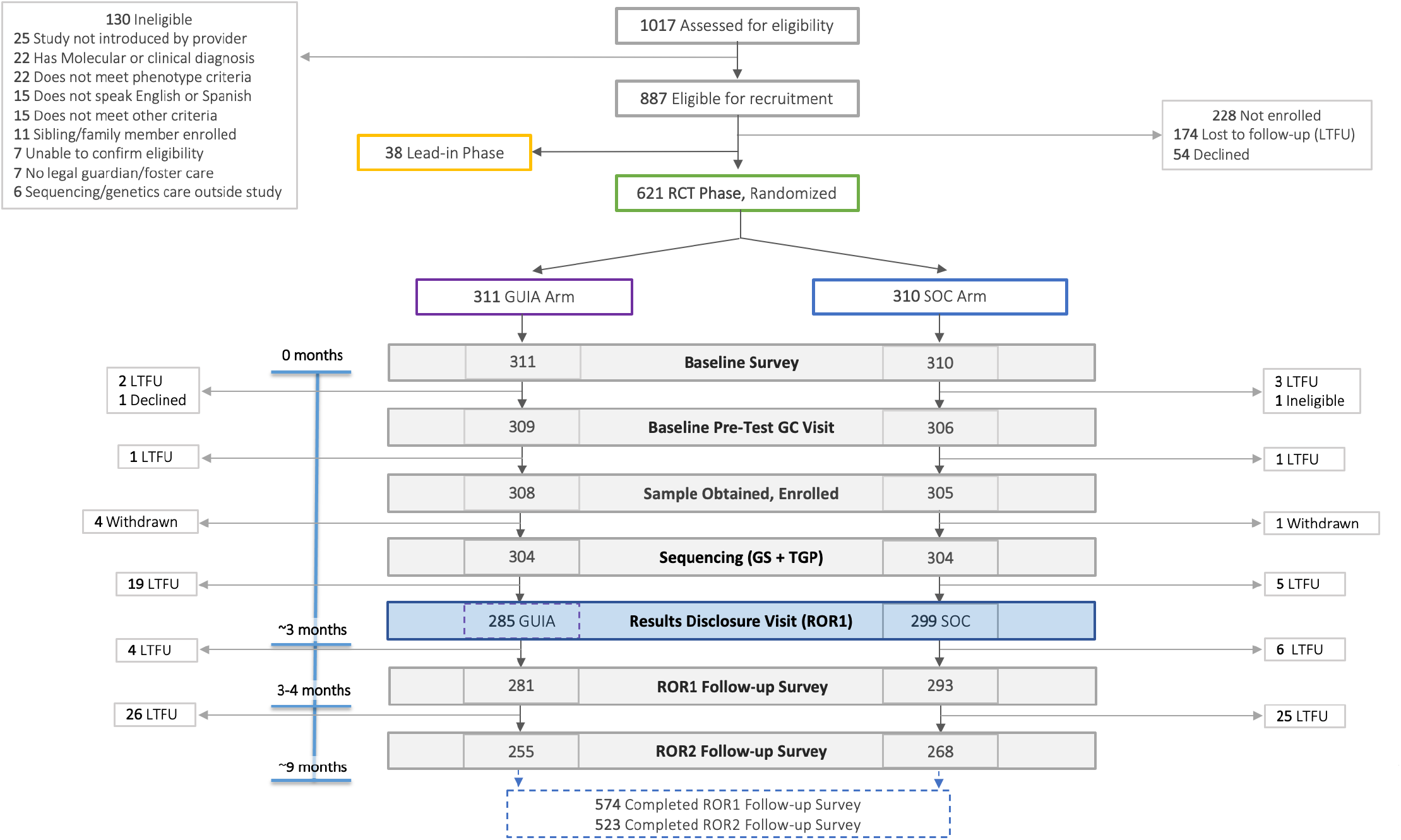
Consort diagram for the NYCKidSeq randomized controlled trial. A total of N=1017 children were assessed for eligibility in NYCKidSeq. Of N=888 who were eligible for enrollment, N=38 were assigned to the Lead-In Phase and N=621 were included in the RCT with N=311 randomized to the GUÍA arm and N=310 to the standard of care (SOC) arm. Excluding the Lead-In Phase (N=37), N=613 enrolled in the RCT; N=308 in the GUÍA arm and N=305 in SOC. In the GUÍA arm, N=281 completed the ROR1 survey and N=255 completed the ROR2 survey. In the SOC arm, N=293 completed the ROR1 survey and N=268 completed the ROR2 survey. Abbreviations: RCT, randomized controlled trial; SOC, standard of care; ROR1, results disclosure time point; ROR2, 6-month post-disclosure time point.

### Study population

The study recruited children with suspected genetic conditions from two healthcare systems in New York City, Mount Sinai Health System (MS) and Albert Einstein College of Medicine/Montefiore Medical Center (EM). For each child, at least one available English- or Spanish-speaking parent or legal guardian (participant) was also recruited to complete study surveys. Children were referred to the study by their healthcare providers at MS or EM. Eligibility criteria included being ≤21 years of age at the time of enrollment and having an undiagnosed neurologic (epilepsy/seizure disorder [epilepsy] or intellectual developmental disability/global developmental delay [IDD]), cardiac (congenital heart disease, cardiomyopathy, or cardiac arrhythmia), or immunologic (features of primary immunodeficiency) phenotype suspected of having a genetic etiology. Those with previously uninformative GT or who had undergone genetic counseling were eligible after three months had passed. Children were excluded if they had a molecular diagnosis for their primary phenotype, an obvious genetic diagnosis based on clinical features, or a bone marrow transplant. We employed a study-wide definition of diversity based on self-reported race/ethnicity (non-White/European ancestry) and/or living in a Health Resources & Services Administration (HRSA) medically underserved area(23,24).

### Recruitment, Enrollment, and Study Visits

Participants and children were recruited and enrolled in the NYCKidSeq study and completed three study visits: baseline enrollment and survey, results disclosure and survey (ROR1), and 6-month post-disclosure survey (ROR2; see **Figure 1**). The study commenced on January 31, 2019, and was completed on April 28, 2022. Per the CSER consortium guidelines, we initially targeted the enrollment of 1,100 children and their families to the study. However, due to institutionally required changes in clinical research prompted by the COVID-19 pandemic, we amended the enrollment target to 650. Children referred to the study (N=1,017) were assessed for eligibility, and parents/legal guardians of those eligible (N=887) were approached for recruitment. Of these, 228 were either lost to follow-up or declined, and 659 expressed interest in participating. The first 38 consented children and participants were assigned to the study’s Lead-in Phase, and data collected throughout their study visits was used to improve and refine the surveys and GUÍA.(21) Data from the Lead-in Phase is not used in any downstream analysis. The subsequent 621 children and participants were consented to the RCT and randomized to either the control or intervention arm using the randomization module in REDCap applied by a study staff member. The stratified randomization scheme balanced primary phenotype (neurologic, cardiac, immunologic) and institution (MS, EM) across each arm. All 621 participants completed the baseline consent and survey, 615 completed the baseline pre-test visit, and blood and/or saliva samples were collected from 613 children and their biological parent(s), if available. These 613 children and participants were designated as being successfully enrolled in the study.

Prior to GT, five participants withdrew from the study. For the remaining 608 children from MS (N=386) or EM (N=222), clinical genome sequencing (GS) and targeted gene panels (TGPs) testing were performed at the New York Genome Center and Sema4, respectively. TGPs consisted of a neurodevelopmental panel, immunodeficiency panel, and cardiovascular panel, and one or multiple panel(s) were run based on the child’s phenotype. All genetic testing was New York State approved and Clinical Laboratory Improvement Amendments (CLIA)-certified, and variant interpretation and reporting were performed independently at each of the two laboratories. Study genetic counselors (GCs) reviewed both diagnostic GT report(s) to determine a case-level (clinical) interpretation of the results. Each child was assigned a case-level interpretation of positive, likely positive, uncertain, or negative. More details on diagnostic GT and case-level interpretation in NYCKidSeq are described in Abul-Husn *et al.*(25).

Fourteen participants were lost to follow-up prior to results disclosure. We disclosed results to 584 participants, 574 completed ROR1, and 523 completed ROR2. Overall, 85.3% (523/613) of participants enrolled in the RCT completed the study. Families received an incentive of up to $80 for completing the study visits. We obtained written informed consent from children >18 years who were cognitively able, from parents/legal guardians of children <18 years or who were unable to consent themselves, and from parents/legal guardians providing samples and/or completing the study surveys. Further details on recruitment, enrollment, and inclusion/exclusion criteria are described in Odgis *et al*.(23).

### Genetic Counseling in Standard of Care (Control) and GUÍA (Intervention) Arms

Since standard of care (SOC) genetic counseling is not well defined(26), we modeled SOC genetic counseling on clinical pediatric genetics settings for this study. Pre-test counseling, which included education, informed consent prior to GT, and family history assessments, was conducted the same way for both arms. The components of the post-test results disclosure session included reviewing the purpose of the GT, disclosure of the GT results, clinical implications of the results, including medical and support referrals when appropriate, disclosure of any secondary findings, family implications, and the provision of relevant resources. In both arms, GCs used a Spanish-speaking medical interpreter in disclosure sessions for participants with limited English proficiency (19.7%; 115/584). Pre- and post-test genetic counseling visits occurred in person until the COVID-19 pandemic in March 2022, which prompted a rapid transition to telehealth. GCs in the SOC arm utilized visual aids at their discretion, which were displayed to the family via screen-sharing during telehealth visits. The GCs provided families with a copy of the child’s GT results, and families could be given additional resources, educational handouts, and/or results summary materials at the GC’s discretion.

In the intervention arm, GCs mimicked SOC and utilized GUÍA for all aspects of the results disclosure. Prior to the visit, GCs entered the child’s results and pertinent result-related information into GUÍA’s purpose-built, web-based GC form, which contained fields for primary and secondary results that included variants and genes, inheritance pattern, description of the condition, affected organ and/or systems, and hyperlinks to resources. The form also contained fields for report personalization, including child’s name, pronouns, and preferred language. GUÍA’s patient-facing, personalized, webpages were then generated, containing digital education modules, the children’ results, family implications, and details on medical recommendations and next steps. Prior to the result disclosure visit, the GC sent a copy of the GUÍA report to the referring provider for their review and input. Participants and GCs worked together to navigate the different sections of the GUÍA display during the session, which was shown on iPad/computer monitor for in-person counseling, or via screen-sharing for telehealth visits. GCs displayed the Spanish version of GUÍA at their discretion (35.1%; 100/285). After the session, a PDF or printout of the full content of GUÍA was provided to the family as well as a copy of the child’s GT results. For both arms, GCs documented the results disclosure visit and uploaded the GT results to the child’s electronic health record (EHR). A PDF of the GUÍA report was also uploaded to the EHR for those in the intervention arm. All visit documentation was routed to the referring provider.

### Outcome Measures and Surveys

Outcomes, demographics, and other participant and child characteristics were collected using surveys administered by trained study staff at baseline, ROR1, and ROR2. The primary outcomes of the RCT are participants’ perceived understanding of their child’s GT results and their perceived confidence in explaining the results to others in the intervention arm vs. the control arm at ROR1 and ROR2. We assessed the primary outcomes using two novel questions. The first asked participants to rate their understanding of the GT results on a scale of 1 (’very little or none of it’) to 5 (’almost all or all of it’), and the second to rate their confidence in explaining their child’s genetic test results to someone else on a scale of 1 (’not confident at all’) to 5 (’completely confident’) (see **Supplemental Methods Table 1**).

The secondary outcome is the participants’ objective understanding of the child’s GT results in the intervention arm vs. the control arm at ROR1 and ROR2. Objective understanding was measured using four novel questions asked of participants. After completing the results disclosure visit, GCs also answered these questions (see **Supplemental Methods Table 1**). We measured the agreement between GCs and participants by matching the participant’s responses to the GC’s. We did not evaluate the additional secondary outcomes of participants’ understanding of the actionability of the results and adherence to medical follow-up recommendations due to inconsistencies in the administration of the survey questions and differences in participants’ interpretation and responses to the survey questions (see **Supplemental Methods**). **Supplemental Table 1** contains all outcome measures and survey time points.

We collected information on a wide range of characteristics of participants and children, including age, biological sex, education, type of insurance, household income, and health literacy, using survey instruments that were novel or adapted from previously validated scales, many of which were harmonized across the CSER consortium(23,27). Participant race and ethnicity were collected using the CSER harmonized self-reported measure that originated from the 2020 Census recommendation(27). This single-item measure combined race and ethnicity, allowing selection from nine choices: seven racial and/or ethnic groups, prefer not to answer, or unknown/none of these describe me. For this analysis, we defined the following population groups: Hispanic/Latino(a) (H/L) ethnicity was prioritized such that participants who selected H/L were re-categorized as H/L regardless of any other race designation made. Participants that selected more than one race were re-categorized into “More than one race”. All other population groups remained if they were the only selection made by the participant. We also collected information on cohort characteristics and activities during study visits, including the language the survey was administered in (language), health system, and the case-level interpretation of the results (see **Supplemental Table 2**). Study staff inputted all survey measurements into a REDCap database.

### Analysis

The primary analytic population consists of randomized participants who completed the baseline survey, attended the results disclosure ROR1 visit, and completed the ROR1 survey within four weeks of results disclosure (N=551). A ROR2 analytic sample used the same criteria above, plus completing the ROR2 survey and not receiving an amended result that changed the clinical interpretation between ROR1 and ROR2 surveys (N=487; see **Supplemental Methods**). We used descriptive statistics to explore outcome frequencies of baseline, ROR1, and ROR2 survey data. We assessed equivalency between the two arms using the chi-square or Fisher’s exact test for categorical variables and the Wilcoxon rank sum or t-tests for continuous variables. The following covariates were included in all models: parent age, education level, language, child’s insurance, case-level interpretation. GC was also included as a covariate in models for primary outcomes, and health system was used instead of GC for the secondary outcome.

We used ordinal logistic regression to evaluate between arm differences for perceived understanding and confidence (primary outcomes) and logistic regression for objective understanding (secondary outcome). We generated an objective understanding summary score by summing the number of questions with an agreement between the GC and participant for the four binary objective understanding questions. The summary score ranged from 0-4, with a higher number indicating better objective understanding.

We performed longitudinal (repeated measure) analysis using ROR1 and ROR2 data to calculate the cumulative odds ratios via generalized estimating equations. We analyzed the primary outcomes using alternating logistic regression with a fully exchangeable working correlation structure. For the secondary outcome, we used a binomial distribution with a logit link for the individual objective understanding questions and a Poisson distribution with a log link for the summary score. We repeated the longitudinal analysis, stratified by health literacy levels (adequate (N=351) and inadequate/marginal (N=200), case-level interpretation (positive/likely positive (N=91), uncertain (N=317) and negative (N=143), the three largest neurological phenotypes IDD (N=183), epilepsy (N=151) and both (N=179)), the three largest population groups (Hispanic/Latino(a) (H/L; N=246), Black/African American (AA; N=84), and White/European American (EA; N=130)), poverty level (at or below 200% of the New York City federal poverty level (N=230) and above (N=258), and interpreter use (with interpreter (N=107) and without interpreter (N=444). The H/L group (N=246) was further stratified by the use of an interpreter at results disclosure (with interpreter (N=104) and without interpreter (N=142)) and by health literacy level ((adequate (N=138) and inadequate/marginal (N=108)). All statistical analyses were performed using SAS software version 9.4 (SAS, Cary, North Carolina).

## Results

### Participant Characteristics

Of the N=608 families enrolled in the RCT, N=551 parents/legal guardians (participants) completed both the baseline and ROR1 survey, attended the results disclosure visit, and were thus included in the analysis. **Table 1** displays the characteristics of all N=551 participants, with N=281 in the SOC arm and N=270 in the GUÍA arm. Participants were asked which racial and/or ethnic category(ies) best described them, and responses were transformed into population groups, prioritizing H/L ethnicity, as described in the Methods section. Population groups were only available for biological parents (N=531), where 46.3% self-reported as H/L, 24.5% as EA, 15.8% as AA, 7.2% Asian, 1.5% Middle Eastern/North African/Mediterranean, 0.4% American Indian/Native American/Alaska Native, 1.3% more than one population, 0.7% other, 1.1% unknown/none of these describe me, and 1.1% preferred not to answer. Participants’ mean age was 41.1 years (range 20.9-81.5), 88.6% were mothers, and 62.3% reported having less than a college degree. 53.9% of participants lived in a medically underserved area, 41.8% were at or below 200% of the New York City federal poverty level, and 64.8% of children had public insurance. Additionally, 36.3% of participants had inadequate or marginal health literacy, and 19.4% used a medical interpreter during results disclosure. Most children (90.9%) had a primary neurological phenotype. 12.9% of children received positive results, 3.6% received likely positive results, 25.9% negative, and 57.5% uncertain. There was equivalence across study arms for all baseline covariates (**Table 1**) except case-level interpretation (*P*=0.003); the SOC arm included more children with positive and negative results, while there were more children with uncertain results in the GUÍA arm. All analytical models included case-level interpretation as a covariate to control for this variance.

**Table 1.**
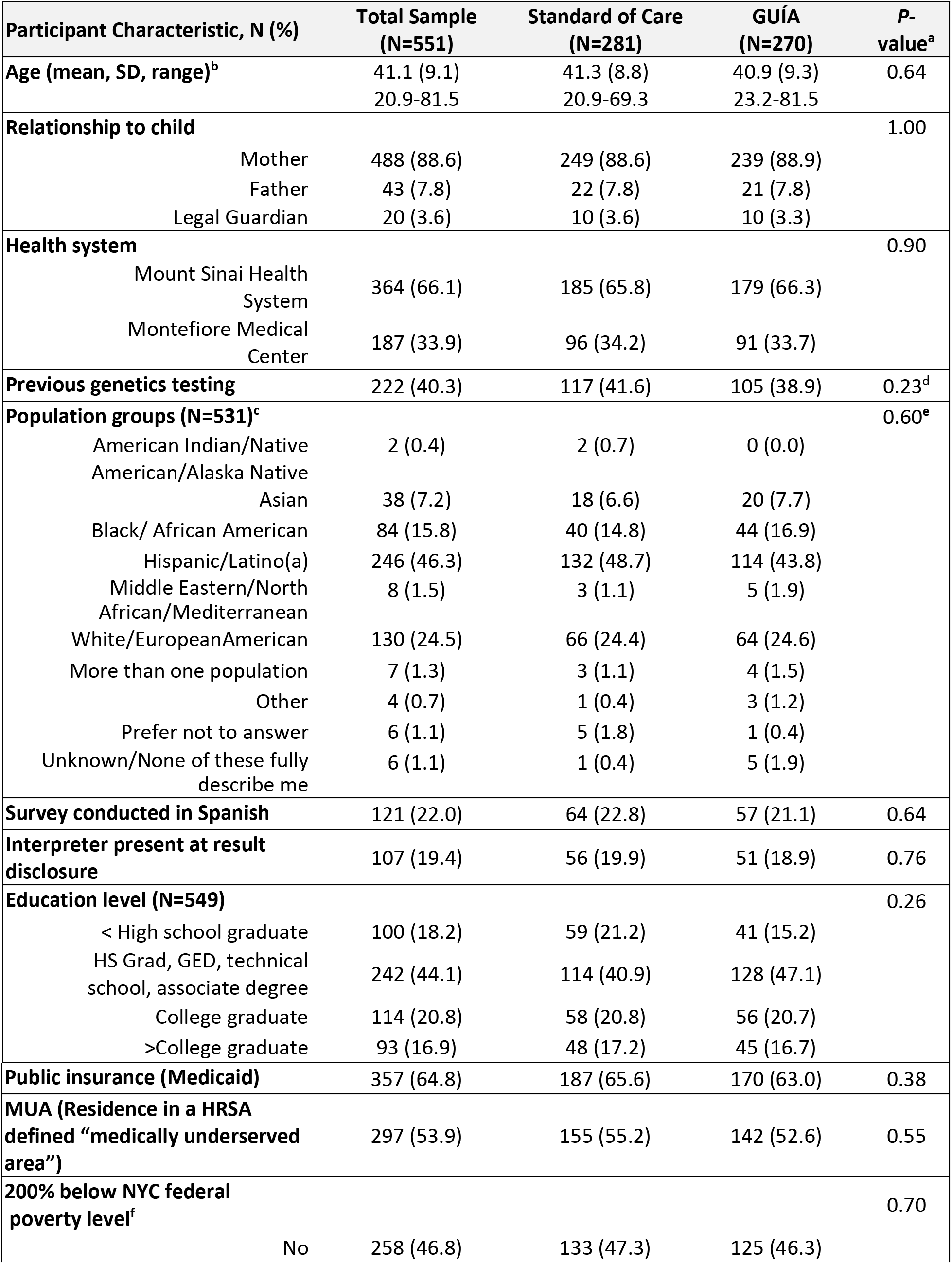
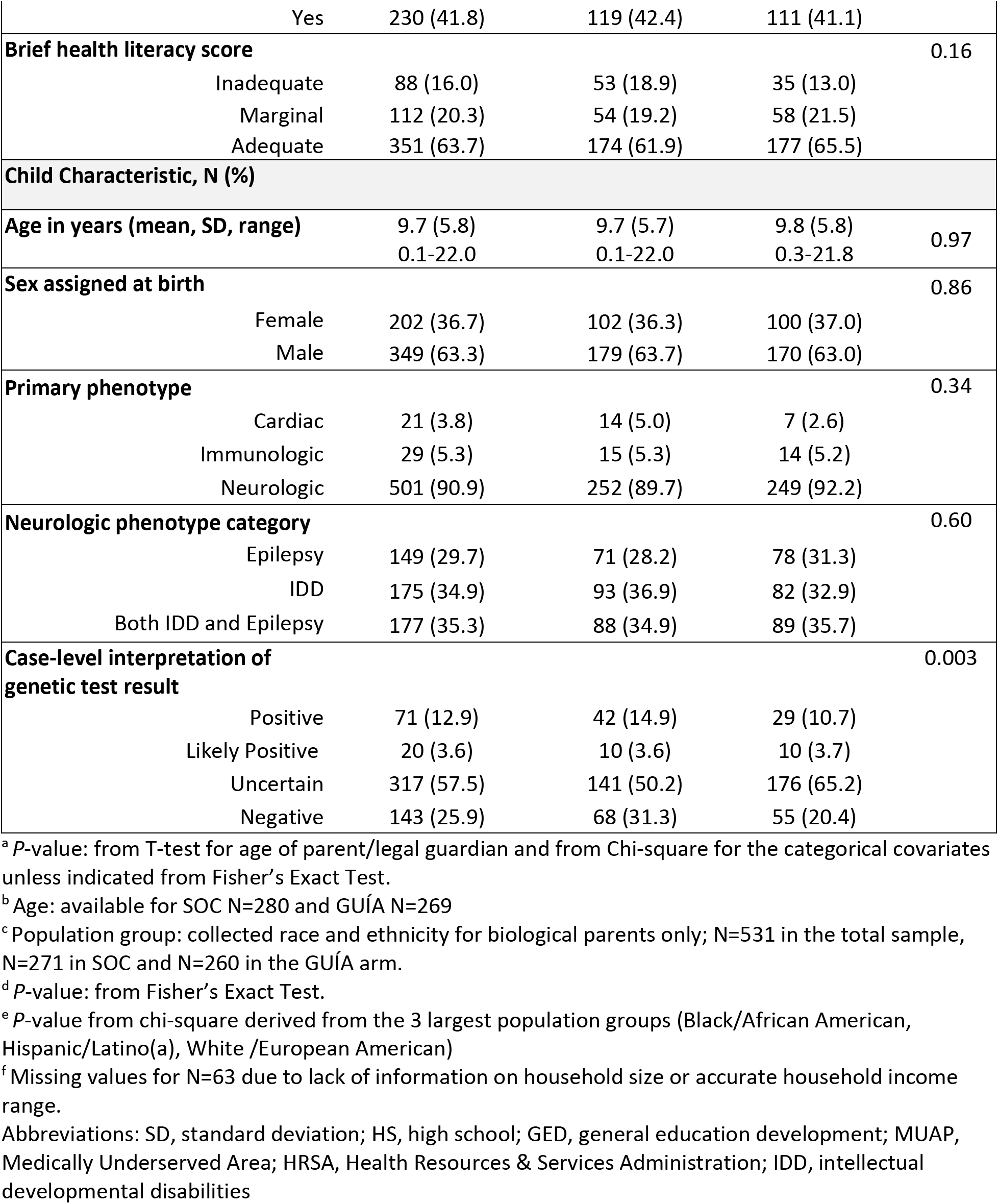
Participant and child characteristics for the total sample and by study arm.

### Participants’ Overall Understanding of Genetic Test Results

We used three measures to evaluate participants’ understanding of their child’s GT results: perceived understanding, perceived confidence in explaining the results to others, and objective understanding. Overall, most participants (58.8%, N=324) selected the highest level of understanding (level 5 ‘almost all or all of it’) following results disclosure (ROR1), and 45.6% (N=196) selected this level approximately 6 months after results disclosure (ROR2). Additionally, at ROR1, 45.6% (N=251) of participants reported the highest level of confidence in their ability to explain the GT results to others, and 33.7% (N=163) selected this level at ROR2. For objective understanding, there was over 69% agreement between the participant and GC for each question at ROR1 and ROR2. The mean objective understanding summary score (range 0 - 4) for the cohort was 2.9 (standard deviation [SD]=1.2) at ROR1 and 2.8 (SD=1.3) at ROR2. We evaluated which covariates of those we measured impacted overall understanding of GT results. Education level, insurance type, population group, GC, health system, case-level interpretation, and survey language were significantly associated with one or both primary and secondary outcomes (see **Supplemental Table 3**) and were included as covariates in downstream analyses.

### Impact of GUÍA on Understanding

The primary study outcome was the impact of the GUÍA intervention on participants’ perceived understanding of and confidence in explaining the GT results up to four weeks (ROR1) and approximately 6 months (ROR2) after receiving results. We first assessed the impact of GUÍA on participants’ perceived understanding at the two time points (ROR1 and ROR2) separately. In a fully adjusted model for perceived understanding at ROR1, participants in the GUÍA arm were more likely to choose the highest level of understanding (level 5) than the lower levels compared to those in the SOC arm (odds ratio [OR]=2.8, 95% confidence interval [CI] 1.004,7.617, *P*=0.049). No significant differences were identified between the arms for perceived understanding at ROR2. Additionally, there were no significant differences between the arms for perceived confidence at ROR1 or ROR2, comparing the highest level (level 5) to the lower levels. The study’s secondary outcome was GUÍA’s ability to improve participants’ objective understanding of the GT result. The objective understanding summary score was not significantly associated with the study arms at ROR1 or ROR2.

We next assessed the impact of GUÍA on participants’ understanding over time by conducting a repeated measure analysis across the ROR1 and ROR2 time points. We did not observe a significant difference between the arms for perceived understanding and confidence (**Figures 2a and 2b**). However, participants in the GUÍA arm maintained a higher objective understanding summary score than those in SOC (OR=1.1, CI[1.004, 1.127], *P*=0.038; **Figure 2c**). The results from this analysis did not change when education level was removed from the models, indicating that education level is not driving the observed difference in objective understanding. Together, these findings suggest that GUÍA was effective in helping participants maintain their objective understanding of their child’s genomic test results over time; however, across the two time points, it did not impact their perceived understanding of the GT results or their confidence in explaining their child’s results to others.

**Figure 2.**
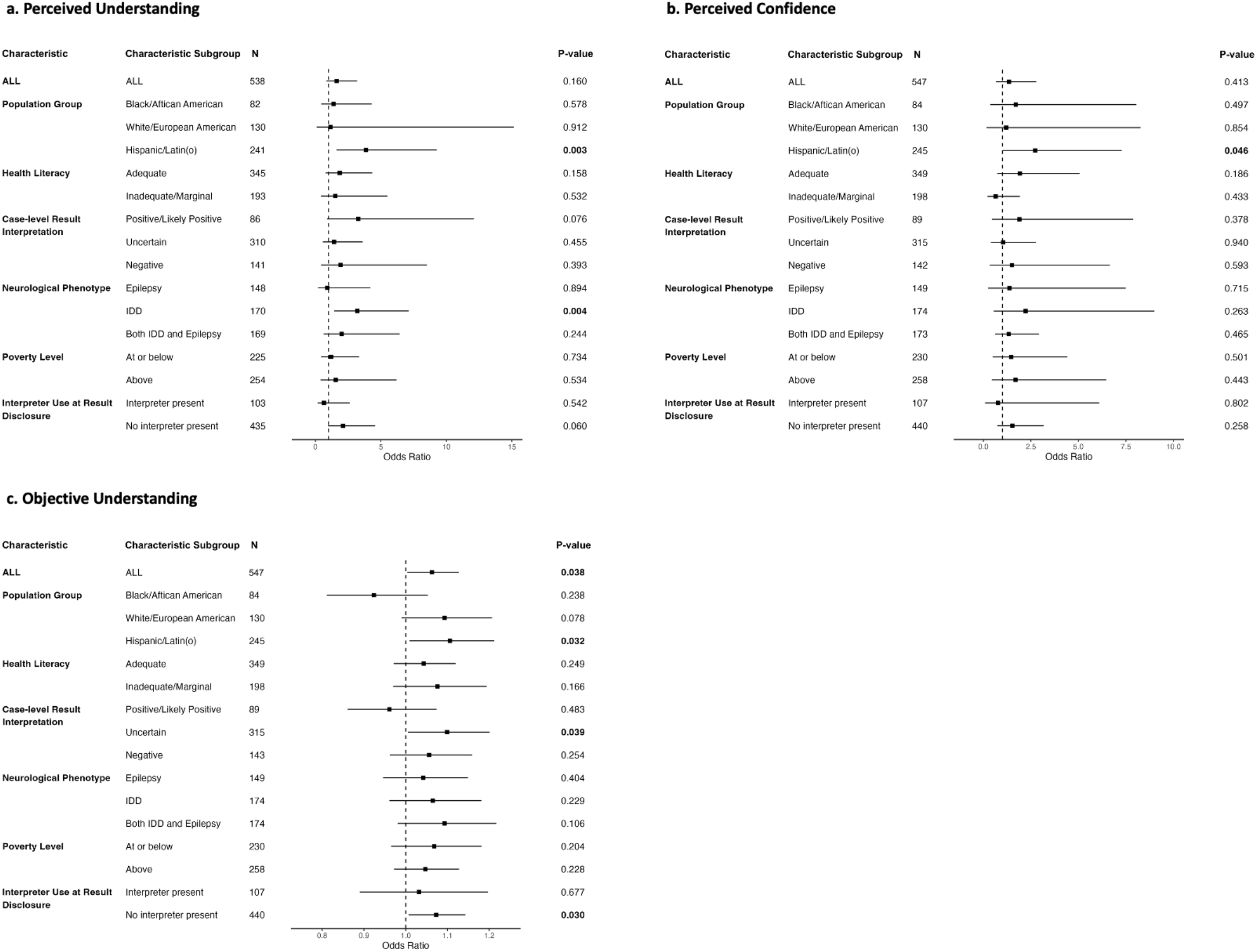
The forest plots display results from the longitudinal analyses using repeated measures assessing the full cohort (All) and stratified into six sub-groups; Population Group, Health Literacy, Case-level Result Interpretation, Neurological Phenotype, Poverty Level, and Interpreter Use at Result Disclosure. Shown in the separate panels are differences in perceived understanding **(a)**, perceived confidence **(b)**, and objective understanding **(c)** between GUÍA and SOC arms for all randomized participants and stratified by characteristic subgroups. We analyzed perceived understanding and confidence using alternating logistic regression, and we used Poisson distribution to analyze the objective understanding summary score. The following covariates were included in all models: controlling for parent age, education level, language, child’s insurance, and case-level interpretation. In addition, genetic counselor was added as a covariate for perceived understanding and confidence and health system was added as a covariate for objective understanding. Abbreviations: SOC, standard of care; H/L, Hispanic/Latino(a); IDD, intellectual developmental disabilities

### Effectiveness of GUÍA across Participant Characteristics

To obtain additional insight into how GUÍA impacts understanding, we performed a stratified analysis of understanding over time considering six socioeconomic and clinical characteristics: health literacy level, poverty level, interpreter use, population group, case-level interpretation, and neurological phenotype. GUÍA was associated with a positive impact on understanding for four specific subgroups (see **Figure 2**). Participants in the GUÍA arm whose children had a neurological phenotype of IDD had higher perceived understanding than those in SOC (OR=3.2, CI[1.4, 7.1], *P*=0.004). Participants whose children received uncertain results maintained a higher objective understanding summary score (OR=1.1, CI[1.005, 1.201], *P*=0.039), as did those who did not use an interpreter (OR=1.1, CI[1.007, 1.143], *P*=0.030). Finally, we observed that H/L participants in the GUÍA arm maintained significantly higher perceived understanding (OR=3.9, CI[1.6, 9.3], *P*=0.003) and confidence (OR=2.7, CI[1.021,7.277], *P*=0.046; see **Figures 2a and 2b**), and maintained a higher objective understanding summary score (OR=1.1, CI[1.009, 1.212], *P*=0.032; **Figure 2c**) compared to SOC. There were no significant differences between the arms for AA and EA population groups for all three measures of understanding.

To further investigate the impact of GUÍA on understanding GT results in the H/L population, we stratified this population by use of a medical interpreter and health literacy level. H/L participants in the GUÍA arm who did not use an interpreter during results disclosure maintained higher perceived understanding and confidence (OR=12.6, CI[3.9, 40.5], p<0.001 and OR=5.4, CI[1.7, 17.8], *P*=0.005, respectively) and a higher objective understanding summary score (OR=1.2, CI[1.035, 1.316], *P*=0.012) (**Figure 3**). However, for H/L participants who used an interpreter, no significant associations with understanding were identified between the arms. H/L participants with adequate health literacy had higher perceived understanding and confidence in the GUÍA arm (OR=7.2, CI[2.3, 22.5], *P*=0.001 and OR=6.0, CI[1.4, 26.4], *P*=0.018, respectively; **Figure 3**). However, there were no significant differences in objective understanding between the arms for H/L participants with inadequate or marginal health literacy. These findings demonstrate that while GUÍA positively impacted H/L participants’ understanding of the results overall, it did not provide a significant benefit for the sub-groups of H/L participants who used a Spanish-speaking interpreter during the results disclosure session or had inadequate or marginal health literacy.

**Figure 3.**
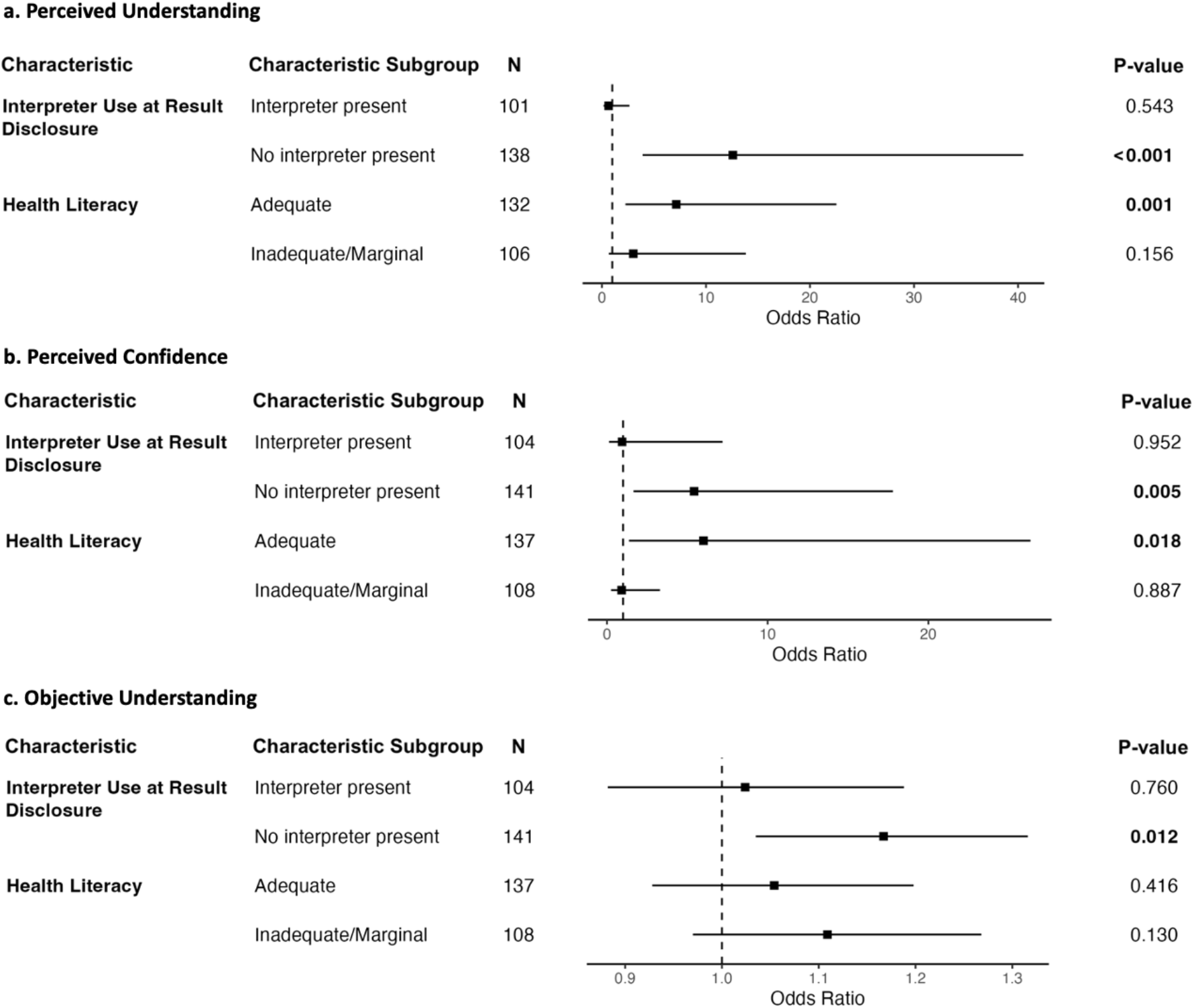
The forest plots display results from the longitudinal stratified analysis using repeated measures of the H/L population by medical interpreter use at result disclosure and health literacy level. The panels display differences in perceived understanding **(a)**, perceived confidence **(b)**, and objective understanding **(c)** between GUÍA and SOC arms for H/L participants stratified by medical interpreter use and health literacy level. We analyzed perceived understanding and confidence using alternating logistic regression, and we used Poisson distribution to analyze the objective understanding summary score. The following covariates were included in all models: controlling for parent age, education level, language, child’s insurance, and case-level interpretation. In addition, genetic counselor was added as a covariate for perceived understanding and confidence and health system was added as a covariate for objective understanding. Abbreviations: SOC, standard of care; H/L, Hispanic/Latino(a); IDD, intellectual developmental disabilities

## Discussion

We evaluated the impact of a novel digital application called GUÍA, designed to facilitate the communication of GT results, on understanding of those results in parents of children with suspected genetic conditions. Families in this study represent diverse, multilingual, and predominantly medically underserved communities in New York City. We showed that GUÍA positively impacted parents’ understanding of their child’s GT results, demonstrating a 2.8-fold increase in perceived understanding immediately following results disclosure and modestly improving their objective understanding by 1.1-fold over time. Stratified analyses provided a more nuanced discernment of GUÍA’s impact, demonstrating that GUÍA significantly increased understanding for parents whose children had a neurological phenotype of IDD and for parents whose children received uncertain GT results. We also demonstrated that the effectiveness of GUÍA in improving understanding was most significant in H/L populations, where we observed a 3-4-fold increase in the primary understanding measures over time. To our knowledge, this is the first study assessing the impact of digitally enhanced genetic counseling to support the delivery of GT results.

Helping patients and their families understand their GT results is an essential aspect of genomic medicine and an important role of genetic counseling(28). Previous work has shown that patients and families can struggle to understand GT results(29), and that results can be misunderstood and misinterpreted by patients, families, and providers, especially when the results are uncertain or have a complicated clinical interpretation(19,30). This is a concern since the level of understanding people have about their results can influence downstream decision-making about medical care, family, and life planning. All these factors make understanding a critical outcome to consider when evaluating novel approaches and interventions for delivering GT results. However, understanding is a complex, multifaceted construct that is challenging to measure, and there are currently no validated or accepted survey instruments designed to assess parents’ understanding of their child’s GT results. For these reasons, we developed three novel measures of understanding to gain a more thorough and nuanced evaluation of parents’ understanding of the GT results. We demonstrated that GUÍA specifically improved objective understanding of uncertain results, typically the most challenging to communicate due to the complex nature of the results, limited available data on the implications of the finding, and ambiguity surrounding the clinical significance and subsequent steps in medical management(20,31,32). We also note a significant impact on understanding in parents of children with IDD, a multifactorial disorder where the underlying etiology can be difficult to determine, suggesting GUÍA may have increased parents’ understanding of results in the context of heightened clinical complexity. In this way, we are directly measuring the effect of a digital tool on one of the essential aspects of genetic counseling encounters, which in turn may help to guide the development of digitally enhanced solutions for scaling genomic medicine.

Almost 80% of parents included in this study were from underrepresented and medically underserved populations. Because GUÍA was specifically designed with diverse communities in mind(21), we evaluated its utility in different population subgroups. A key finding was that GUÍA was most helpful in increasing understanding outcomes for the H/L population in our study. These results suggest that GUÍA may be a useful and beneficial tool for counseling individuals of H/L descent, who currently represent almost 19% of the US population(33) yet for whom research has demonstrated significant unmet needs in genomic medicine(34). Interestingly, GUÍA’s effectiveness in the H/L population was attenuated when a medical interpreter was used for result disclosure. Although there are cited challenges with using a medical interpreter, such as the possibility for interpreters conveying inaccurate information or lacking knowledge of the correct translations of genetic terminology(35,36), we had designed GUÍA to address these challenges by displaying both English and Spanish text on the GUÍA interface. It is possible that the reading level of the Spanish text was still too high or that the participants could not process the written text while listening to an interpreter. Nevertheless, this approach was inadequate in overcoming obstacles associated with a language discordance between families and GCs. Addressing this barrier will be critical in the future for ensuring GUÍA is effective in linguistically diverse populations, where there is already a lack of high-quality genetic counseling resources(37) and where only 1% of the genetic counseling profession speaks a language other than English(38).

There are limitations to this study that should be considered. The NYCKidSeq study was ongoing during the COVID-19 pandemic, which impacted the clinical environment and changed how research was conducted, including shifting from in-person to telehealth genetic counseling. This may have affected parents’ experiences and the study outcomes in ways not measured in this study. Additionally, most children had a neurologic phenotype, which limits our insight into how GUÍA will perform in other clinical contexts. We were interested in whether GUÍA affected parents’ understanding of and adherence to medical recommendations. However, these outcomes were difficult to assess given the complexity of the clinical care for enrolled children and the limitations of the CSER harmonized survey instruments to capture the full spectrum of medical care. Increasing diversity among study participants helps advance our understanding of the clinical utility of genomic medicine in all populations and ensures that genetic research is broadly applicable. However, if there are differences between groups, stratification analysis to uncover those differences reduces statistical power (although reduced environmental variability within a sub-group may also increase power). For example, stratifying by population group uncovered a significant signal in H/L (the largest group), but no significant signal in EA or AA, which could be due to insufficient power in those groups. Also, we excluded several population groups (e.g., Asian) from the stratified analysis due to low sample size. In general, broadening diversity in research studies teaches both about generalizable and specific effects; however, the latter may need to be followed up with further well-powered studies.

## Conclusions

The NYCKidSeq RCT demonstrated that using GUÍA to enhance GT result disclosure digitally positively impacted parents’/legal guardians’ perceived and objective understanding of their child’s GT results, most significantly in H/L populations. These findings illustrate that digital tools can be developed for diverse families with sick children and applied in clinically complex contexts to improve understanding. Future work could also explore additional functionalities and use cases for GUÍA, such as evaluating GUÍA as a tool for non-genetics providers increasingly offering GT in their clinics who often encounter challenges communicating genomic information outside their expertise(39). As advances in deep learning and generative AI increase, burgeoning opportunities exist to continue to augment genetic counseling by providing more interactive and personalized interfaces, chatbot-based communications, and making genomic information more approachable for patients and providers. Research frameworks, such as the one developed for this study, will be vital to assess the effectiveness of digital tools for enhancing the communication of genomic information in myriad clinical or public health settings. In the future, advancing digital technology could help scale genomic medicine and empower patients’ downstream medical decision-making.

## List of abbreviations

AA: Black/African American
AI: Artificial intelligence
CI: 95% confidence interval
CLIA: Clinical Laboratory Improvement Amendments
CSER: Clinical Sequencing Evidence-Generating Research
EA: White/European American
EHR: Electronic health record
EM: Albert Einstein College of Medicine/Montefiore Medical Center
GC: Genetic counselor
GUÍA: Genomic Understanding, Information, and Awareness application
GS: Genome sequencing
GT: Genomic tests
HRSA: Health Resources & Services Administration
H/L: Hispanic/Latino(a)
IDD: Intellectual developmental disability/global developmental delay
MS: Mount Sinai Health System
OR: Odds ratio
RCT: Randomized controlled trial
SOC: Standard of care
TGPs: Targeted gene panels

## Declarations

### Ethics approval and consent to participant

The Institutional Review Boards at the Icahn School of Medicine at Mount Sinai (IRB# 17-00780) and the Albert Einstein College of Medicine (IRB# 2016-6883) approved this study. All participants provided written informed consent. This research conformed to the Declaration of Helsinki.

### Consent for publication

Not applicable

### Availability of data and materials

The datasets used and/or analyzed during the current study are available from the corresponding author upon reasonable request.

### Authors’ contributions

LJB, BDG, CRH, GAD, JMG, MPW, NSAH, REZ, and EEK contributed to the conception and design of the study.

SAS, JAO, KMG, MS, KEB, PNM, KEB, MAR, MDB, and JER contributed to the acquisition of data. LS and BJI performed the analysis.

SAS, NRK, and EEK contributed to the interpretation of data.

SAS, BJI, JAO, KEB, LS, MS, NRK, PNM, and EEK, drafted the manuscript. All authors read and approved the final manuscript.

## Data Availability

The datasets used and/or analyzed during the current study are available from the corresponding authors upon reasonable request.

## Acknowledgements

The authors thank all the children, parents, and families who participated in this study. The authors also thank the New York Genome Center, Rady Children’s Institute for Genomic Medicine, and Sema4 laboratory; the members of the Mount Sinai Hospital Genomics Stakeholder Board; referring physicians in the Mount Sinai and Montefiore Health Systems.

## Additional File 1

**Supplemental Table 1.** Outcomes assessed through the NYCKidSeq study

**Supplemental Table 2.** Covariates and population characteristics included in the analysis

**Supplemental Table 3.** Association of covariates to perceived understanding, confidence, and objective understanding at ROR1

**Supplemental Methods**

## References

1. Linn AJ, van Dijk L, Smit EG, Jansen J, van Weert JCM. May you never forget what is worth remembering: the relation between recall of medical information and medication adherence in patients with inflammatory bowel disease. J Crohns Colitis. 2013 Dec;7(11):e543–50.

2. Dimatteo MR. The role of effective communication with children and their families in fostering adherence to pediatric regimens. Patient Educ Couns. 2004 Dec;55(3):339–44.

3. Patel HV, Henrikson NB, Ralston JD, Leppig K, Scrol A, Jarvik GP, et al. Implementation matters: How patient experiences differ when genetic counseling accompanies the return of genetic variants of uncertain significance. AMIA Annu Symp Proc. 2021;2021:950–8.

4. Hoskovec JM, Bennett RL, Carey ME, DaVanzo JE, Dougherty M, Hahn SE, et al. Projecting the Supply and Demand for Certified Genetic Counselors: a Workforce Study. J Genet Couns. 2018 Feb;27(1):16–20.

5. Gordon ES, Babu D, Laney DA. The future is now: Technology’s impact on the practice of genetic counseling. Am J Med Genet C Semin Med Genet. 2018 Mar;178(1):15–23.

6. Snir M, Nazareth S, Simmons E, Hayward L, Ashcraft K, Bristow SL, et al. Democratizing genomics: Leveraging software to make genetics an integral part of routine care. Am J Med Genet C Semin Med Genet. 2021 Mar;187(1):14–27.

7. Wakefield CE, Meiser B, Homewood J, Taylor A, Gleeson M, Williams R, et al. A randomized trial of a breast/ovarian cancer genetic testing decision aid used as a communication aid during genetic counseling. Psychooncology. 2008 Aug;17(8):844–54.

8. Adam S, Birch PH, Coe RR, Bansback N, Jones AL, Connolly MB, et al. Assessing an Interactive Online Tool to Support Parents’ Genomic Testing Decisions. J Genet Couns [Internet]. 2018 Jul 23; Available from: http://dx.doi.org/10.1007/s10897-018-0281-1

9. Schwartz MD, Valdimarsdottir HB, DeMarco TA, Peshkin BN, Lawrence W, Rispoli J, et al. Randomized trial of a decision aid for BRCA1/BRCA2 mutation carriers: impact on measures of decision making and satisfaction. Health Psychol. 2009 Jan;28(1):11–9.

10. Bombard Y, Clausen M, Shickh S, Mighton C, Casalino S, Kim THM, et al. Effectiveness of the Genomics ADvISER decision aid for the selection of secondary findings from genomic sequencing: a randomized clinical trial. Genet Med. 2020 Apr;22(4):727–35.

11. Green MJ, Peterson SK, Baker MW, Friedman LC, Harper GR, Rubinstein WS, et al. Use of an educational computer program before genetic counseling for breast cancer susceptibility: effects on duration and content of counseling sessions. Genet Med. 2005 Apr;7(4):221–9.

12. Hernan R, Cho MT, Wilson AL, Ahimaz P, Au C, Berger SM, et al. Impact of patient education videos on genetic counseling outcomes after exome sequencing. Patient Educ Couns. 2020 Jan;103(1):127–35.

13. Sanderson SC, Suckiel SA, Zweig M, Bottinger EP, Jabs EW, Richardson LD. Development and preliminary evaluation of an online educational video about whole-genome sequencing for research participants, patients, and the general public. Genet Med. 2016 May;18(5):501–12.

14. Schmidlen T, Schwartz M, DiLoreto K, Kirchner HL, Sturm AC. Patient assessment of chatbots for the scalable delivery of genetic counseling. J Genet Couns. 2019 Dec;28(6):1166–77.

15. Ponathil A, Ozkan F, Welch B, Bertrand J, Chalil Madathil K. Family health history collected by virtual conversational agents: An empirical study to investigate the efficacy of this approach. J Genet Couns. 2020 Dec;29(6):1081–92.

16. Nazareth S, Hayward L, Simmons E, Snir M, Hatchell KE, Rojahn S, et al. Hereditary Cancer Risk Using a Genetic Chatbot Before Routine Care Visits. Obstet Gynecol. 2021 Dec 1;138(6):860–70.

17. Siglen E, Vetti HH, Lunde ABF, Hatlebrekke TA, Strømsvik N, Hamang A, et al. Ask Rosa - The making of a digital genetic conversation tool, a chatbot, about hereditary breast and ovarian cancer. Patient Educ Couns. 2022 Jun;105(6):1488–94.

18. Lee W, Shickh S, Assamad D, Luca S, Clausen M, Somerville C, et al. Patient-facing digital tools for delivering genetic services: a systematic review. J Med Genet. 2023 Jan;60(1):1– 10.

19. Donohue KE, Gooch C, Katz A, Wakelee J, Slavotinek A, Korf BR. Pitfalls and challenges in genetic test interpretation: An exploration of genetic professionals experience with interpretation of results. Clin Genet. 2021 May;99(5):638–49.

20. Suckiel SA, O’Daniel JM, Donohue KE, Gallagher KM, Gilmore MJ, Hendon LG, et al. Genomic Sequencing Results Disclosure in Diverse and Medically Underserved Populations: Themes, Challenges, and Strategies from the CSER Consortium. J Pers Med [Internet]. 2021 Mar 13;11(3). Available from: http://dx.doi.org/10.3390/jpm11030202

21. Suckiel SA, Odgis JA, Gallagher KM, Rodriguez JE, Watnick D, Bertier G, et al. GUÍA: a digital platform to facilitate result disclosure in genetic counseling. Genet Med. 2021 May;23(5):942–9.

22. Amendola LM, Berg JS, Horowitz CR, Angelo F, Bensen JT, Biesecker BB, et al. The Clinical Sequencing Evidence-Generating Research Consortium: Integrating Genomic Sequencing in Diverse and Medically Underserved Populations. Am J Hum Genet. 2018 Sep 6;103(3):319– 27.

23. Odgis JA, Gallagher KM, Suckiel SA, Donohue KE, Ramos MA, Kelly NR, et al. The NYCKidSeq project: study protocol for a randomized controlled trial incorporating genomics into the clinical care of diverse New York City children. Trials. 2021 Jan 14;22(1):56.

24. Find Shortage Areas by Address [Internet]. [cited 2023 Apr 2]. Available from: https://data.hrsa.gov/tools/shortage-area/by-address

25. Abul-Husn NS, Marathe PN, Kelly NR, Bonini KE, Sebastin M, Odgis JA, et al. Molecular diagnostic yield of genome sequencing versus targeted gene panel testing in racially and ethnically diverse pediatric patients. medRxiv [Internet]. 2023 Mar 20; Available from: http://dx.doi.org/10.1101/2023.03.18.23286992

26. Biesecker BB, Lillie SE, Amendola LM, Donohue KE, East KM, Foreman AKM, et al. A review and definition of “usual care” in genetic counseling trials to standardize use in research. J Genet Couns. 2021 Feb;30(1):42–50.

27. Goddard KAB, Angelo FAN, Ackerman SL, Berg JS, Biesecker BB, Danila MI, et al. Lessons learned about harmonizing survey measures for the CSER consortium. J Clin Transl Sci. 2020 Apr 24;4(6):537–46.

28. National Society of Genetic Counselors’ Definition Task Force, Resta R, Biesecker BB, Bennett RL, Blum S, Hahn SE, et al. A new definition of Genetic Counseling: National Society of Genetic Counselors’ Task Force report. J Genet Couns. 2006 Apr;15(2):77–83.

29. Watnick D, Odgis JA, Suckiel SA, Gallagher KM, Teitelman N, Donohue KE, et al. “Is that something that should concern me?”: a qualitative exploration of parent understanding of their child’s genomic test results. HGG Adv [Internet]. 2021 Apr 8;2(2). Available from: http://dx.doi.org/10.1016/j.xhgg.2021.100027

30. Wynn J, Ottman R, Duong J, Wilson AL, Ahimaz P, Martinez J, et al. Diagnostic exome sequencing in children: A survey of parental understanding, experience and psychological impact. Clin Genet. 2018 May;93(5):1039–48.

31. Makhnoon S, Shirts BH, Bowen DJ. Patients’ perspectives of variants of uncertain significance and strategies for uncertainty management. J Genet Couns. 2019 Apr;28(2):313–25.

32. Levin Fridman A, Raz A, Timmermans S, Shkedi-Rafid S. Views of Israeli healthcare professionals regarding communication of genetic variants of uncertain significance to patients. J Genet Couns. 2022 Aug;31(4):912–21.

33. Office of Minority Health [Internet]. [cited 2023 May 2]. Available from: https://minorityhealth.hhs.gov/omh/browse.aspx?lvl=3&lvlid=64#:∼:text=According%20to%202020%20Census%20data,the%20largest%20at%2061.6%20percent.

34. Dron HA, Bucio D, Young JL, Tabor HK, Cho MK. Latinx attitudes, barriers, and experiences with genetic counseling and testing: A systematic review. J Genet Couns. 2023 Feb;32(1):166–81.

35. Joseph G, Lindberg NM, Guerra C, Hernandez C, Karliner LS, Gilmore MJ, et al. Medical interpreter-mediated genetic counseling for Spanish preferring adults at risk for a hereditary cancer syndrome. J Genet Couns [Internet]. 2023 Mar 20; Available from: http://dx.doi.org/10.1002/jgc4.1695

36. Gutierrez AM, Robinson JO, Statham EE, Scollon S, Bergstrom KL, Slashinski MJ, et al. Portero versus portador: Spanish interpretation of genomic terminology during whole exome sequencing results disclosure. Per Med. 2017 Nov;14(6):503–14.

37. Beauchesne R, Birch P, GenCOUNSEL Study, Elliott AM. Genetic counselling resources in non-english languages: A scoping review. PEC Innov. 2023 Dec;2:100135.

38. NSGC > Policy, Research and Publications > Professional Status Survey [Internet]. [cited 2023 May 30]. Available from: https://www.nsgc.org/Policy-Research-and-Publications/Professional-Status-Survey

39. Arora NS, Davis JK, Kirby C, McGuire AL, Green RC, Blumenthal-Barby JS, et al. Communication challenges for nongeneticist physicians relaying clinical genomic results. Per Med. 2016 Sep;14(5):423–31.

