## Supplemental File for "The NYCKidSeq randomized controlled trial: Impact of GUÍA digitally enhanced genetic counseling in racially and ethnically diverse families"

**Additional File 1**

Current affiliations

a. Graduate Program in Human Genetics, Sarah Lawrence College, Bronxville, NY, USA
b. Illumina, San Diego, CA, USA
c. iECURE Incorporated, Philadelphia, PA, USA
d. 23andMe, Inc., Sunnyvale, California, USA

***Corresponding authors:**
Eimear E. Kenny, MSc, PhD
Icahn School of Medicine at Mount Sinai
One Gustave L. Levy Place, Box 1003
New York, NY 10029


Sabrina A. Suckiel, MS
Icahn School of Medicine at Mount Sinai
One Gustave L. Levy Place, Box 1003
New York, NY 10029


### Supplemental Table 1. Outcomes assessed through the NYCKidSeq study

| Measures | Survey question | Response options | Source | Time point | | |
|---|---|---|---|---|---|---|
| | | | | BL | ROR 1 | ROR 2 |
| **Primary Outcomes** | | | | | | |
| Perceived understanding | Genetic test results can be complicated. How much did you understand about the results that were given to you? Please rate your understanding on a scale of 1 to 5, where 1 means "very little or none of it" and 5 means "you understood almost all or all of it." | 1, 1 very little or none of it \| 2, 2 \| 3, 3 \| 4, 4 \| 5, 5 you understood almost all or all of it \| 98, Refused | NYCKidSeq (novel) | - | X | X |
| Perceived confidence | If you needed to explain your child's genetic test results to someone else, how confident would you feel doing so? Answer on a scale of 1 to 5, where 1 is "not confident at all" and 5 is "completely confident. " | 1, 1 not confident at all \| 2, 2 \| 3, 3 \| 4, 4 \| 5, 5 completely confident \| 98, Refused | NYCKidSeq (novel) | - | X | X |
| **Secondary Outcome** | | | | | | |
| Objective Understanding | 1. The result of the genetic test means my child's condition is definitely caused by something in his/her genes.<br>2. The result of the genetic test gave me a genetic explanation for my child's condition/symptoms.<br>3. At this time, there is no genetic explanation for my child's condition/symptoms.<br>4. The result of the genetic test means we still don't know if my child's condition is genetic or not. | 1, Yes \| 0, No \| 97, Not sure/don't know \| 98, Refused | NYCKidSeq (novel) | - | X | X |
| GC Objective Understanding | 1. The result of the genetic test means the child's condition is definitely caused by something in his/her genes.<br>2. The result of the genetic test gave me a genetic explanation for the child's condition/symptoms.<br>3. At this time, there is no genetic explanation for the child's condition/symptoms.<br>4. The result of the genetic test means we still don't know if the child's condition is genetic or not. | 1, Yes \| 0, No \| 97, Not sure/clear \| 98, Refused | NYCKidSeq (novel) | - | X | - |

| | | | | | |
|---|---|---|---|---|---|
| Understanding of medical follow up & actionability 1 | Were any recommendations made based on the test result? | 1, Yes \| 0, No \| 97, I don't know/don't remember \| 98, Refused | Adapted from CSER (novel): Recommended Medical Actions and Follow Through on Recommendations Attributable to Genomic Testing (MRA) | | X |
| Understanding of medical follow up & actionability 2 | [IF YES] What were the recommendations? Check all that apply. | 1, Medication \| 2, Additional non-genomic medical tests for screening, monitoring, or diagnosis (e.g., blood test, imaging such as x-ray, MRI, etc) \| 3, Referrals to consult with other doctors or specialist \| 4, Referral to a non-MD health professional \| 5, Referral for mental health support \| 6, Referral for therapeutic services \| 7, Other | Adapted from CSER (novel): Recommended Medical Actions and Follow Through on Recommendations Attributable to Genomic Testing (MRA) | | X |
| GC anchor - Understanding of medical follow up & actionability 1 | Were any recommendations made based on the test result? | 1, Yes \| 2, No \| 3, I don't know/don't remember | Adapted from CSER (novel): Recommended Medical Actions and Follow Through on Recommendations Attributable to Genomic Testing (MRA) | | X |
| GC anchor - Understanding of medical follow up & actionability 2 | [IF YES] What were the recommendations? Please go through each. | 1, Medication \| 2, Additional non-genomic medical tests for screening, monitoring, or diagnosis (e.g., blood test, imaging such as x-ray, MRI, etc) \| 3, Referrals to consult with other doctors or specialist \| 4, Referral | Adapted from CSER (novel): Recommended Medical Actions and Follow Through on Recommendatio | | X |

| | | | | | | |
|---|---|---|---|---|---|---|
| | | to a non-MD health professional \| 5, Referral for mental health support \| 6, Referral for therapeutic services \| 7, Lifestyle changes | ns Attributable to Genomic Testing (MRA) | | | |
| Adherence to medical follow up recommendations 1 | Did you discuss your child's genetic test results with your child's doctors or health care providers? | 1, Yes \| 2, Not yet but I plan to \| 0, No and I don't plan to \| 98, Refused | CSER (novel): Recommended Medical Actions and Follow Through on Recommendations Attributable to Genomic Testing (MRA) | | | X |
| Adherence to medical follow up recommendations 2 | [IF YES] Did the doctor or health care provider make any recommendations based on the test result? | 1, Yes \| 0, No \| 97, I don't know/don't remember \| 98, Refused | CSER (novel): Recommended Medical Actions and Follow Through on Recommendations Attributable to Genomic Testing (MRA) | | | X |
| Adherence to medical follow up recommendations 3 | [IF YES] What were the recommendations? | 1, Medication \| 2, Additional non-genomic medical tests for screening, monitoring, or diagnosis (e.g., blood test, imaging such as x-ray, MRI, etc) \| 3, Referrals to consult with other doctors or specialist \| 4, Referral to a non-MD health professional \| 5, Referral for mental health support \| 6, Referral for therapeutic services \| 7, Lifestyle changes | CSER (novel): Recommended Medical Actions and Follow Through on Recommendations Attributable to Genomic Testing (MRA) | | | X |

Note: NYCKidSeq measures were developed specifically for this research study. CSER measures were developed by a collaborative group of CSER investigators, as outlined in Goddard et al., 2020(1)

### Supplemental Table 2. Covariates and population characteristics included in the analysis

| Covariate/ Population Characteristic | Survey question | Response options | Source | Time point | | |
|---|---|---|---|---|---|---|
| | | | | BL | ROR 1 | ROR2 |
| Primary parent age | What is your date of birth? | MM/DD/YYYY | NA | X | - | - |
| Relationship to Child | What is your relationship to [child]? | 1, Mother \| 2, Father \| 3, Legal Guardian | NA | X | X | X |
| Biological sex of child | What sex was your child assigned at birth, on the original birth certificate? (Check one) | 1, Female \| 2, Male \| 97, Prefer not to answer | CSER measure adapted from GenIUSS(2) | X | X | X |
| Educational level | What is the highest grade or level of school you completed or the highest degree you received? (Check one) | 1, No schooling completed \| 2, Elementary school (kindergarten through 5th grade) \| 3, Middle school (6th, 7th, or 8th grade) \| 4, Some high school (9th, 10th, or 11th grade) \| 5, 12th grade, no diploma \| 6, High school graduate (diploma or GED or equivalent) \| 7, Some post-high school training (college or occupational, technical, or vocational training), no degree or certificate \| 8, Completed occupational, technical, or vocational program, received degree or certificate \| 9, Associate (2-year) college degree \| 10, Bachelor's degree (for example: BA, AB, BS) \| 11, Master's degree (for example: MA, MS, MEng, MEd, MSW, MBA) \| 12, Professional degree (for example: MD, DDS, DVM, LLB, JD) \| 13, Doctoral degree (for example: PhD, EdD) \| 14, Don't Know \| 98, Prefer not to answer | CSER (novel) | X | - | - |

| | | | | | | |
|---|---|---|---|---|---|---|
| Self-reported race and ethnicity | What category or categories best describe the child's mother [father]? Check all that apply. | 1, American Indian, Native American, or Alaska Native \| 2, Asian \| 3, Black or African American \| 4, Native Hawaiian/Pacific Islander \| 5, White or European American \| 6, Middle Eastern or North African/Mediterranean \| 7, Hispanic/Latino(a) \| 8, Other \| 97, Prefer not to answer \|98, Unknown/none of these fully describe the child's mother [father] | CSER measure adapted from US Census(3) | X | - | - |
| Child's insurance status | Is your child covered by health insurance or some other kind of health care plan? (Include health insurance obtained through employment or purchased directly as well as government programs like Medicare and Medicaid that provide medical care or help pay medical bills) (Check one) | 1, Yes \| 0, No \| 2, Don't Know \| 98, Prefer not to answer | CSER measure adapted from NHANES(4) | X | - | - |
| Type of Insurance | IF YOUR CHILD IS COVERED: What kind or kinds of health insurance or health care coverage does your child have? (Check all that apply) | 1, Private health insurance, employment based \| 2, Private health insurance, directly purchased \| 3, Government plan, Medicare \| 4, Government plan, Medicaid \| 5, Government plan, Military health care \| 6, Other type of insurance \| 0, No coverage of any type \| 7, Refused | CSER measure adapted from NHANES(4) | X | - | - |
| Household income | What was your household's total family income (before taxes) from all sources in the last year? (Check one) | 0, Less than $5,000 \| 1, $5,000 to $9,999 (monthly $416 to $833, biweekly $208 to $416) \| 2, $10,000 to $14,999 (monthly $833 to $1,249, biweekly $416 to $624) \| 3, $15,000 to $19,999 ($1,249 to $1,666 , $624 to $833) \| 4, $20,000 to $24,999 ($1,666 to $2,083 , $833 to $1,041) \| 5, $25,000 to $29,999 ($2,083 to $2,499 , $1,041 to $1,249) \| 6, $30,000 to $39,999 ($2,499 to $3,333 , $1,249 to $1,666) \| 7, $40,000 to $49,999 ($3,333 to $4,166 , $1,666 to $2,083) \| 8, $50,000 to $59,999 ($4,166 to $4,999 , $2,083 to $2,499) \| 9, $60,000 to $69,999 ($4,999 to $5,833 , $2,499 to $2,916) \| 10, $70,000 to | CSER measure adapted from NHANES(4) | X | - | - |

| | | | | | | |
|---|---|---|---|---|---|---|
| | | $79,999 ($5,833 to $6,666 , $2,916 to $3,333) \| 11, $80,000 to $99,999 ($6,666 to $8,333 , $3,333 to $4,166) \| 12, $100,000 to $119,999 ($8,333 to $9,999 , $4,166 to $4,999) \| 13, $120,000 to $139,999 ($9,999 to $11,666 , $4,999 to $5,833) \| 14, $140,000 or more ($11,666+ , $5,833+) \| 16, Don't Know \| 15, Prefer not to answer | | | | |
| Number of people supported | How many people (children and adults) were supported by this income in the last year? | Text | CSER measure adapted from NHANES(4) | X | - | - |
| Prior exposure to genetic testing | Has anyone in your family (including yourself and your child) ever had a genetic test? | 1, Yes \| 0, No \| 97, Don't Know \| 98, Prefer not to answer | NYCKidSeq (novel) adapted from Genetic testing to Understand and Address Renal Disease Disparities (GUARDD) study(5) | X | - | - |
| Health literacy 1 | How often do you have someone (like a family member, friend, hospital/clinic worker or caregiver) help you read medical materials? | 1, Always \| 2, Often \| 3, Sometimes \| 4, Occasionally \| 5, Never \| 6, Refused | CSER measure adapted from BRIEF Health Literacy Survey(6) | X | - | - |
| Health literacy 2 | How often do you have problems learning about your medical conditions because of difficulty understanding written information? | 1, Always \| 2, Often \| 3, Sometimes \| 4, Occasionally \| 5, Never \| 6, Refused | | X | - | - |
| Health literacy 3 | How often do you have a problem understanding what is told to you about your medical condition? | 1, Always \| 2, Often \| 3, Sometimes \| 4, Occasionally \| 5, Never \| 6, Refused | | X | - | - |
| Health literacy 4 | How confident are you in filling out medical forms by yourself? | 1, Not at all \| 2, A little bit \| 3, Somewhat \| 4, Quite a bit \| 5, Extremely \| 6, Refused | | X | - | - |
| **Covariates and population characteristics collected by study staff** | | | | | | |
| Group assignment | Randomization | 1, Traditional GC \| 2, Communication Tool | NA | X | - | - |

| | | | | | | |
|---|---|---|---|---|---|---|
| Medically underserved area | Is this address located in a HRSA defined 'medically underserved area' (MUA)? | 0, No \| 1, Yes | HRSA MUA(7) | X | - | - |
| Date of survey administration | Date baseline [ROR1, ROR2] survey was administered | MM/DD/YY | NA | X | X | X |
| Language survey administered | What language is this survey being administered? | 1, English \| 2, Spanish | NA | X | X | X |
| Child’s age | Referred Child's DOB | MM/DD/YYYY | NA | X | - | - |
| Health system | Hospital / Health System | 1, Montefiore \| 2, Mount Sinai | NA | X | - | - |
| Primary indication | Primary Indication - Disease category | 1, Cardiac \| 2, Immunologic \| 3, Neurologic | NA | X | - | - |
| Neurologic phenotype | Neurologic Disease Category | 1, Epilepsy \| 2, IDD | NA | X | - | - |
| Case-level interpretation | Clinical interpretation Do not include incidental or ACMG 59 findings. | 1, Positive \| 2, Likely positive \| 3, Uncertain \| 4, Negative | NA | - | X | - |
| Amended clinical interpretation | Amended clinical interpretation | 1, Positive \| 2, Likely positive \| 3, Uncertain \| 4, Negative | NA | - | X | - |
| Disclose amended report | Did you disclose an amended report finding to the patient? | 0, No \| 1, Yes | NA | - | X | - |
| Timing of amended results disclosure | When did you disclose the amended results? | 1, At ROR1 visit \| 2, After ROR1 and before ROR2 \| 3, After ROR2 | NA | - | X | - |
| Date of results disclosure | Date of post-test visit | MM/DD/YYYY | NA | - | X | - |
| Genetic counselor | Who returned the results? | Names of study genetic counselors (N=8) | NA | - | X | - |
| Present at results disclosure | Who was present at the post-test GC visit? Select all that apply. | 1, Child's (biological) mother \| 2, Child's (biological) father \| 3, Child's legal guardian(s) \| 4, Child (proband) \| 5, Other | NA | - | X | - |
| Interpreter use | Was an interpreter used for this visit? | 0, No \| 1, Yes | NA | - | X | - |

**Supplemental Table 3. Association of covariates to perceived understanding, confidence, and objective understanding at ROR1**

| | Perceived Understanding | | | Perceived Confidence | | | Objective Understanding[a] | | |
|---|---|---|---|---|---|---|---|---|---|
| **Covariate** | **N** | **OR (95% CI) or Wald Chi-Square[b]** | ***P* value** | **N** | **OR (95% CI) or Wald Chi-Square[b]** | ***P* value** | **N** | **OR (95% CI) or Wald Chi-Square[b]** | ***P* value** |
| **Health System** *Mount Sinai vs Montefiore Medical Center* | 542 | 1.5 (1.0, 2.1) | **0.04** | 551 | 1.2 (0.9, 1.7) | 0.26 | 551 | 1.0 (0.9, 1.1) | 0.65 |
| **Genetic Counselor[c]** *N=8 categories* | 542 | 16.9 | **0.02** | 551 | 27.7 | 0.15 | 551 | 5.9 | 0.55 |
| **Age[d]** | 540 | 1.0 (1.0, 1.0) | 0.97 | 549 | 1.0 (1.0, 1.0) | 0.55 | 549 | 1.0 (1.0, 1.0) | 0.34 |
| **Education Level** *N=3 categories* | 540 | 13.5 | **0.004** | 549 | 19.7 | **<0.001** | 549 | 4.1 | 0.25 |
| **Survey Language** *English vs Spanish* | 542 | 1.0 (0.7, 1.5) | 0.90 | 551 | 0.9 (0.6, 1.4) | 0.71 | 551 | 1.1 (1.0, 1.3) | **0.05** |
| **Population Groups[e]** *N=3 categories* | 454 | 20.9 | **<0.001** | 460 | 8.4 | **0.02** | 460 | 11.3 | **0.004** |
| **Insurance Type** *Medicaid vs Non-Medicaid* | 542 | 0.6 (0.4, 0.9) | **0.007** | 551 | 0.6 (0.4, 0.8) | **0.002** | 551 | 0.9 (0.8, 1.0) | **0.004** |
| **Case-level Interpretation of Results[f]** *N=4 categories* | 542 | 3.7 | 0.29 | 551 | 5.7 | 0.14 | 551 | 30.3 | **<0.001** |

a. The objective understanding summary score was used for the analysis.
b. Wald Chi-Square was run for categorical variables with 3 or more categories.
c. Eight genetic counselors provided result disclosure genetic counseling to participants.
d. Participant age was collected at baseline.
e. Population groups included the three largest population groups (Black or African American, Hispanic/Latino(a), White or European American).
f. Case-level interpretation of results included positive, likely positive, uncertain, or negative categories.

Abbreviations: OR, odds ratio; CI, confidence interval; ROR1, results disclosure time point

### Supplemental Methods

**Data Extraction and Cleaning**

Study data was collected and managed using REDCap (Research Electronic Data Capture), a secure web-based software platform hosted at the Icahn School of Medicine at Mount Sinai. Data collection instruments captured study activities and participant engagement, including but not limited to referral eligibility, recruitment, randomization, enrollment, consent and data sharing preferences, sample acquisition, study procedures, genetic findings, and survey assessments collected by clinical research coordinators and genetic counselors.

Data cleaning was conducted systematically throughout the project with final cleaning and data processing following the completion of study-specific time points, and at the end of data collection in May 2022. Data was queried for potential entry errors, missingness, outliers, and other quality assurance and control inquisitions ensuring the integrity of the data. Study staff reviewed queries and checked source material including REDCap instruments and audit logs, chart notes in the electronic medical record at pre-test and post-test, and paper versions of the phenotype checklists, testing requisition forms, genetic reports, and participant surveys. If the REDCap entry and source material did not match, the source material was used to reconcile the data. If the REDCap entry was missing, the source material was checked to determine if the value was an entry error or if it was a true missing. If the value was an input error, and verified by source, changes were made to the raw data in REDCap. If the value was truly missing, the entry would remain as such. If the REDCap entry matched the source material but was a clear error based on QA across another variable, decisions were made on how to handle these discrepancies. All queries, final resolutions and those that remained unresolved, and changes to the data were recorded in audit and change logs. Upon resolution of all data queries, raw data was exported from REDCap for analysis.

**Participant Inclusions and Exclusions**

The primary inclusion criterion was randomization to either GUIA or SOC arm; participants assigned to the lead-in feedback phase were excluded (N=37). Among the randomized subjects, the primary parent/legal guardian must have completed the baseline survey (to capture baseline characteristics and potential confounders), attended the post-test GC visit, and completed the ROR1 survey to be eligible. Additionally, participants who completed the ROR1 survey more than four weeks after receipt of their genetic test results were ineligible (N=1). Applying this exclusion and inclusion criteria yielded the ROR1 analytic sample of 551 participants.

A ROR2 analytic sample was subsequently constructed. The eligible criteria included participation in the ROR1 analytic sample and completion of the ROR2 survey. Moreover, if an amended genetic report was returned between ROR1 and ROR2 that changed the clinical interpretation and required additional counseling, they were excluded from the ROR2 analytic sample (N=1). Of the 551 participants in the ROR1 analytic sample, 487 are included in the ROR2 analytic sample due to participants lost to follow-up (N=52), changes in primary parent (N=11) and amended reports that changed the clinical interpretation between ROR1 and ROR2 (N=1).

### Primary Outcome Measures

*Perceived understanding and confidence*

Participants' perceived understanding of their child's genomic test results was assessed using a novel survey question on the ROR1 and ROR2 surveys (see **Supplemental Table 1** for question and response options). Response options were transformed for regression analysis of ROR1 data, the lowest response level (1) was omitted due to power (N=9) and levels 2 and 3 were combined, producing three analytic levels (2/3, 4, and 5). For consistency, ROR2 response options were similarly transformed resulting in individuals responding with level 1 being omitted (N=21). Missing data was not included in the final analysis (N=1 at ROR2).

Perceived confidence in explaining genetic test results was measured using a novel question asked in the ROR1 and ROR2 surveys (see **Supplemental Table 1** for question and response options). There were no analytic transformations of the data, and missing data (N=3 at ROR2) was excluded from the final analysis. Partial proportional regression models were run controlling for genetic counselor, parental education, parental age, language at ROR1/ROR2 survey, child's insurance, and clinical interpretation.

Ordinal logistic regression was used to evaluate between arm differences in perceived understanding and confidence. When a covariate in the regression model did not satisfy the proportional odds assumption, the partial proportional odds model was used adjusting for GC, parent age (continuous), education level, language of ROR1/ROR2 surveys, child's insurance type, and case-level interpretation of the results.

### Secondary Outcomes Measures

*Objective Understanding*

The secondary outcome of participants' objective understanding of the child's GT results was measured using four novel questions designed for this study and asked of participants in the ROR1 and ROR2 surveys. The genetic counselor (GC) who conducted the result disclosure visit answered the same questions upon completion of the visit (see **Supplemental Table 1** for questions and response options). GCs' responses were reviewed and mapped to clinical interpretation, creating a standardized "correct responses" set for the individual questions. For example, the correct responses for a positive or likely positive result were "yes" to questions 1 and 2 and no to questions 3 and 4. GC responses were compared to this mapping, and discrepancies were queried. A GC reviewed the queries against the results, post-test note, and GUÍA (if applicable) to confirm findings and how the results were communicated to the family. The clinical team and group decisions reviewed the response to queries that were made: 1) change original response to standardized response (N=58 cases), or 2) leave original response and note edge case due to case complexity/nuance (N=4). A new variable was created to capture the responses that were changed. For analysis, the participant's response was transformed by comparing their response to the GC's for each question; if the responses were the same, it was coded as 'match; if it differed, it was coded as 'no match,' producing a binary variable.

Logistic regression was conducted to analyze the four binary objective understanding variables, controlling for clinical interpretation, health system, parent's age, education, and insurance status. We did not control for genetic counselors since they were not associated with objective understanding. A summary objective understanding score was calculated as the sum of the number of matches from the four binary objective understanding questions (ranging from 0 to 4) with a higher number indicating better objective understanding. The summary score was analyzed using Poisson regression to account for the count nature of the variable. Missing data was not included in the final analysis (N=1 at ROR2).

*Actionability of results and adherence to medical follow-up recommendations*

Understanding of the actionability of genomic results was collected after results disclosure (ROR1) using an adapted CSER (novel) measure ("Recommended Medical Actions and Follow Through on Recommendations Attributable to Genomic Testing (MRA)"), and adherence to medical follow-up recommendations was asked at ROR2 using the CSER (novel) MRA measure (see **Supplemental Table 1** for questions and response options). Due to inconsistencies in how the questions were asked between the ROR1 and ROR2 surveys, and differences in how participants interpreted and answered the survey questions, we were unable to evaluate and report on these outcomes.

**Transformation of covariates and select characteristics of interest**

*Parent Age:* Age of the parent/legal guardian was collected at baseline **(Supplemental Table 2)** and calculated by subtracting the participant's self-reported data of birth from the date of baseline survey administration and was used as a continuous variable for analysis. Missing values were not included in the final analysis (N=2).

*Education Level:* Education level of the parent was collected at baseline (**Supplemental Table 2)**. For analysis, education level was collapsed into four categories: less than high school graduate (response options 1-5), high school graduate/GED, technical school, or associate degree (response options 6-9), college graduate (option 10), and college graduate plus (options 11-13). Those that selected don't know or prefer not to answer were not included in the final analysis (N=2).

*Child's Insurance:* Insurance status of the child was collected on the baseline survey (**Supplemental Table 2**). For analysis, insurance status was binarized into "Public", which included any Government plan, and "Private", which included the private health insurance and other plans, if not government.

*Population Groups:* Race and ethnicity was collected at baseline (**Supplemental Table 2**). For population characteristics and analysis, Hispanic/Latino(a) ethnicity was prioritized; participants who selected Hispanic/Latino(a) were re-categorized as Hispanic/Latino(a) regardless of any other race designation made. Participants that selected more than one race were re-categorized into "More than one race". All other race and ethnicity categories remained if they were the only selection made by the participant. Due to power, the three largest race and ethnicity groups were accessed in stratification analyses: Hispanic/Latino(a) (H/L), White or European American (EA), and Black or African American (AA). Race and ethnicity was not collected for legal guardians (N=20) and were therefore excluded from the analysis.

*Household Income and Number of People Supported:* Poverty index was calculated using participant reported family income and number of people supported collected at baseline **(Supplemental Table 2).** If mean household income, accounting for the number of people supported, was at or below 200% of the 2022-2023 NYC Federal Income Guidelines, participants were categorized as living in poverty. Missing responses, 'don't know' or 'prefer not to answer' were excluded (N=63). Additionally, one participant with a reported income of '$140,000 or more' that supported 15 people was excluded as we were unable to determine whether they fell below or above the poverty level. However, 91 participants reported >$140,000 with a household size that ranged from 2 to 8 and were classified as above poverty level.

*Health Literacy Level:* Health literacy was captured at baseline using four survey items (**Supplemental Table 2).** Response options were summed in order to create the health literacy summary score and categorized by range of score: inadequate (4-12), marginal (13-16), and adequate (17-20). Per CSER analysis guidelines, mean imputations were calculated for those that did not provide responses for no more than half of the health literacy items (N=1).

*Health System:* Study research coordinators indicated the health system (MS or EM) from which participants were recruited at referral **(Supplemental Table 2).**

*Genetic Counselors:* Eight study genetic counselors from MS and EM were assigned to one of the study arms; those involved in the development of GUÍA were assigned to the GUIA arm (N=4) and the remaining assigned to the SOC arm (N=4) (**Supplemental Table 2).**

*Survey Language Administration:* The language (Spanish or English) in which surveys were administered was documented in the study database by study staff **(Supplemental Table 2)**. Survey language at ROR1 was used in all analyses using ROR1 data and survey language at ROR2 was used in all analyses using ROR2 data. For repeated measures analyses, language at each appropriate time point was used in the model.

*Medical Interpreter Use:* Genetic counselors documented whether a Spanish interpreter was used during the result disclosure visit **(Supplemental Table 2).**

*Case-Level Interpretation of Genetic Test Results:* Clinical interpretation of GT results was categorized by the genetic counselors as positive, likely positive, uncertain or negative based on criteria previously described in Abul-Husn et al. 2023.(8) An interpretation committee made up of study physicians with a background in medical genetics was created if discrepancies were found between the genome sequencing and targeted gene panel results. For stratified analyses, positive and likely positive were collapsed due to power, while negative and uncertain remained as they were.

*Primary Indication for Testing and Neurologic Phenotype Category*: The primary indication for testing was collected at recruitment from the referring provider using a study specific phenotype checklist. Providers indicated if primary concern was neurologic, cardiac, or immunologic. Participants were randomized on this selection. Participants could have more than one indication for testing. Each indication category had additional phenotype terms for further characterization. Neurologic: if epilepsy and/or intellectual developmental disability/global developmental delay; Cardiac: if congenital heart disease, cardiomyopathy, and/or cardiac arrhythmia; and Immunologic: features of immunodeficiency. Primary indication testing data did not undergo any transformations and no data was missing. Neurologic phenotype category was analyzed for those with a primary indication of Neurologic. Due to power, further phenotype characterization of those with a primary cardiac and immunologic indication were not analyzed.

**References**

1. Goddard KAB, Angelo FAN, Ackerman SL, Berg JS, Biesecker BB, Danila MI, et al. Lessons learned about harmonizing survey measures for the CSER consortium. J Clin Transl Sci. 2020 Apr 24;4(6):537–46.

2. Williams Institute (University of California, Los Angeles. School of Law). Best Practices for Asking Questions to Identify Transgender and Other Gender Minority Respondents on Population-based Surveys. 2014.

3. Website [Internet]. Available from: Jones NA. Update on the U.S. Census Bureau’s Race and Ethic Research for the 2020 Census. Survey News 2015;3(5). (https://www.census.gov/content/dam/Census/newsroom/press-kits/2014/article_race_ethnic_research_2020census_jones.pdf

4. 1999-2000 Questionnaire Data - Continuous NHANES [Internet]. [cited 2023 Jun 2]. Available from: https://wwwn.cdc.gov/nchs/nhanes/search/DataPage.aspx?Component=Questionnaire&CycleBeginYear=1999

5. Horowitz CR, Abul-Husn NS, Ellis S, Ramos MA, Negron R, Suprun M, et al. Determining the effects and challenges of incorporating genetic testing into primary care management of hypertensive patients with African ancestry. Contemp Clin Trials. 2016 Mar;47:101–8.

6. Haun, J., Noland Dodd, V. J., Graham-Pole, J., Rienzo, B., & Donaldson, P. (2009). Testing a Health Literacy Screening Tool: Implications for Utilization of a BRIEF Health Literacy Indicator. Federal Practitioner, 26(12), 24-31

7. MUA Find [Internet]. [cited 2023 Jun 2]. Available from: https://data.hrsa.gov/tools/shortage-area/mua-find

8. Abul-Husn NS, Marathe PN, Kelly NR, Bonini KE, Sebastin M, Odgis JA, et al. Molecular diagnostic yield of genome sequencing versus targeted gene panel testing in racially and ethnically diverse pediatric patients. Genet Med. 2023 May 5;100880.
